# Functional proteomic profiling links deficient DNA clearance to mortality in patients with severe COVID-19 pneumonia

**DOI:** 10.1101/2022.01.25.22269616

**Authors:** Iker Valle Aramburu, Dennis Hoving, Spyros I. Vernardis, Martha Tin, Vadim Demichev, Elisa Theresa Helbig, Lena Lippert, Klaus Stahl, Marianna Ioannou, Mia I. Temkin, Matthew White, Helena Radbruch, Jana Ihlow, David Horst, Scott T. Chiesa, John E. Deanfield, Sascha David, Christian Bode, Florian Kurth, Markus Ralser, Venizelos Papayannopoulos

## Abstract

Hyperinflammation, coagulopathy and immune dysfunction are prominent in patients with severe infections. Extracellular chromatin clearance by plasma DNases suppresses such pathologies in mice but whether severe infection interferes with these pathways is unclear. Here, we show that patients with severe SARS-CoV-2 infection or microbial sepsis exhibit low extracellular DNA clearance capacity associated with the release of the DNase inhibitor actin. Unlike naked DNA degradation (DNase), neutrophil extracellular trap degradation (NETase) was insensitive to G-actin, indicating distinct underlying mechanisms. Activity-based proteomic profiling of severely ill SARS-CoV-2 patient plasma revealed that patients with high NETase and DNase activities exhibited 18-fold higher survival compared to patients with low activity proteomic profiles. Remarkably, low DNA clearance capacity was also prominent in healthy individuals with chronic inflammation, suggesting that pre-existing inflammatory conditions may increase the risk for mortality upon infection. Hence, functional proteomic profiling illustrates that non-redundant DNA clearance activities protect critically ill patients from mortality, uncovering protein combinations that can accurately predict mortality in critically ill patients.

## Main

Microbial sepsis and severe SARS-CoV-2 infection cause mortality with few available treatment options^1, 2, 3^. These conditions share common pathological features such as hyper-inflammation, coagulopathy, and immune dysfunction that can culminate to organ failure^4, 5 6^. Sepsis due to secondary co-infection is also frequent in patients with severe SARS-CoV-2 infection^7^. The mechanisms that counter these pathologies and decrease mortality are poorly understood. Cell-free chromatin can be found in the circulation of microbial sepsis and SARS-Cov-2 patients and has been linked to hyperinflammation, coagulopathy and immune dysfunction in murine models of sepsis^8, 9, 10, 11, 12^. Mice lacking DNase I and DNase I L3, the two DNases present in the circulation, develop severe thrombosis upon sterile and microbial challenge^13^. Moreover, low DNA and NET clearance activity has been associated with chronic autoimmunity and severe thermal injury^14, 15, 16, 17, 18^. However, the impact of acute infection on DNA clearance mechanisms and their contribution to survival remain unknown.

We recently showed that T cell death promotes pathology and mortality in a murine model of fungal sepsis by releasing histones in the circulation^19^. Cell death was substantially decreased in the spleens of T cell deficient mice stained for CD3 and apoptotic chromosomal fragmentation using terminal deoxynucleotidyl transferase dUTP nick end labelling (TUNEL) (**Fig. 1A and S1A**), that exhibited improved survival^19^. To understand the impact of T cell death in disease pathology we employed mass spectrometry to analyse the plasma proteomes of naïve WT and TCR-α deficient mice or mice infected intravenously with *C. albicans*. Infected TCR-α deficient mice exhibited a distinct proteomic profile compared to WT control animals (**Fig. 1B and 1C**). Actin was amongst the plasma proteins that were increased upon infection in WT mice but remained significantly lower in TCR-α deficient mice (**Fig. 1D and S1B**). Elevated actin in the circulation have been reported in patients with microbial sepsis, SARS-CoV-2 infection or severe injury^20, 21, 22, 23, 24^. Monomeric G-actin and filamentous F-actin are potent inhibitors of DNase I, an endonuclease that along with DNase I L3 degrade extracellular DNA and suppress thrombosis^13, 25, 26, 27^. Consistently, major clotting components such as fibrinogen A, B and G (FGA, FGB and FGG) were elevated in WT mice but were decreased in mice lacking T cells (**Fig. 1D and S1B**). These data indicated that T cell death and actin release corelate with an increased risk for thrombosis in this model.

**Figure 1.**
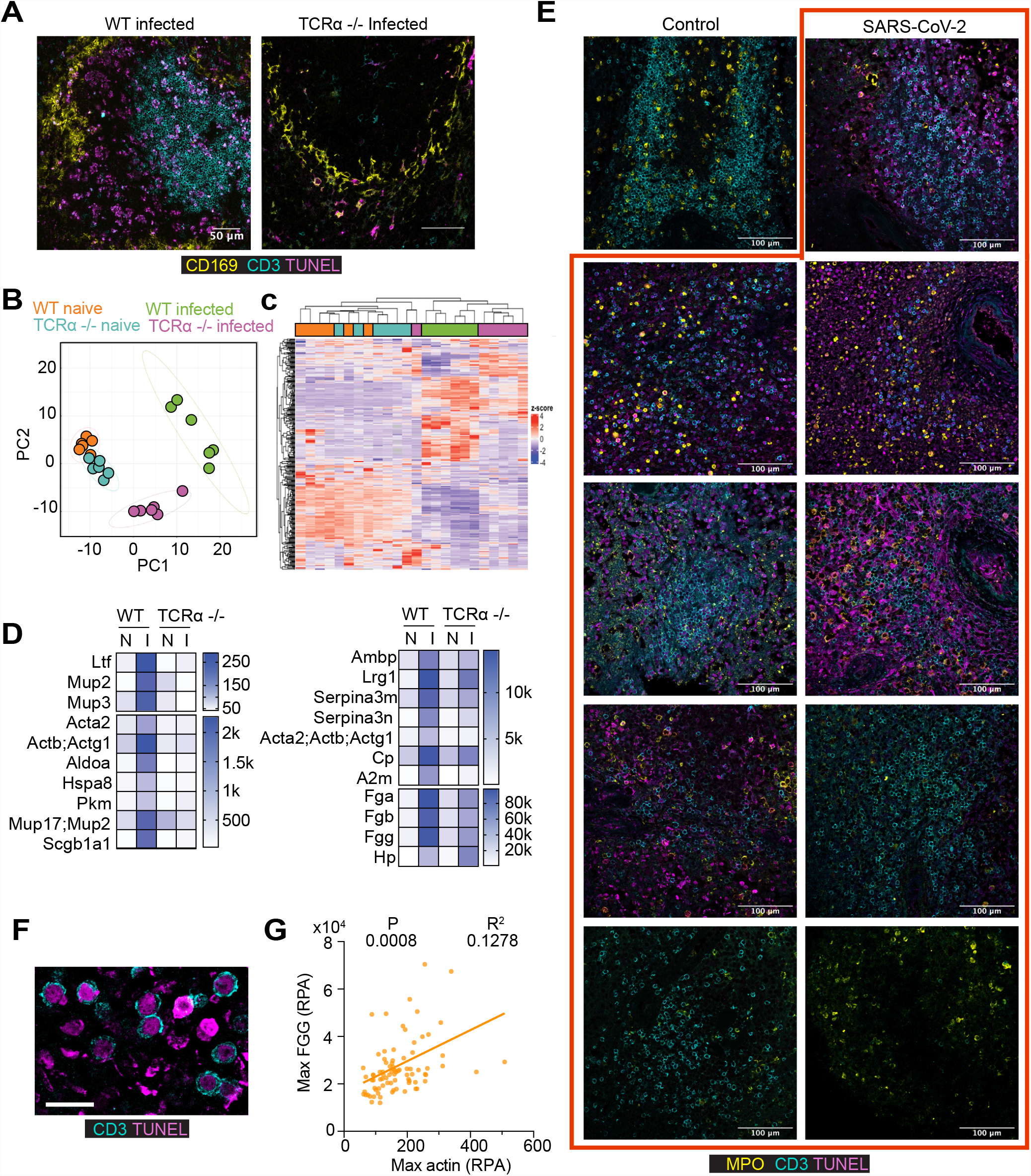
Splenic T cell death promotes actin release and increased risk for coagulopathy. **A**. Immunofluorescence confocal micrographs of spleens from WT and TCRα-deficient mice infected intravenously with *C. albicans*, 3 days post-infection and stained for CD169 (marginal zone macrophages), CD3 (T cells) and TUNEL (apoptosis). Scale bars: 50 μm. **B**. PCA analysis of plasma proteomes from mice in (A) analysed by mass spectrometry. **C**. Heatmap depicting hierarchical clustering by protein expression in the plasma of mice in (A). **D**. Heatmap of proteins that are upregulated in infected WT mice but remain low in infected TCRα-deficient animals. **E**. Immunofluorescence confocal micrographs of post-mortem spleens from a control patient and 9 patients who succumbed to SARS-CoV-2 infection, stained for CD3, TUNEL and myeloperoxidase (MPO, neutrophils). Scale bars: 100 μm. **F**. High magnification micrographs from (E). Scale bars: 20 μm. **G**. Correlation plots between proteomic measurements of the maximum longitudinal actin and fibrinogen G (FGG) levels. Fitting by simple linear regression.

To explore whether such mechanisms are relevant in SARS-CoV-2 infection, we stained post-mortem spleen tissues from patients for T cells, TUNEL and myeloperoxidase (MPO) to document neutrophil infiltration. Extensive splenic T cell death was evident in 6 out the 9 patient samples with T cells comprising a substantial portion of TUNEL+ cells (**Fig. 1E and 1F**). These findings are consistent with previous reports of lymphocytopenia and cell death in the spleens of SARS-CoV-2 infected patients^28, 29^. We performed a correlation analysis of longitudinal plasma proteomes from 63 SARS-CoV-2 patients (CP) with a maximum WHO severity grade 7. The relationship between changes in the plasma proteomes in this cohort and disease severity has been established in previous work^24, 30, 31^. This analysis indicated a significant positive correlation between the maximum actin and FGG levels per patient (**Fig. 1G**). These data suggested that splenocyte death is associated release of cell-free actin in the circulation and an elevated risk for thrombosis.

Given the ability of actin to inhibit DNase I activity, we examined the DNA clearance capacity in the plasma of 36 patients with microbial sepsis (SP) and 43 SARS-CoV-2 infected patients classified with World Health Organization (WHO) ordinal severity scale grades of 3, 4 and 7. We compared the ability of patient and healthy donor (HD) plasmas to degrade plasmid DNA by gel electrophoresis which we termed DNase activity (**Fig. S2A**) and to dissolve neutrophil extracellular traps (NETs), monitored by time-lapse microscopy, which we termed NETase activity (**Fig. S2B)**. We employed both assays because we discovered that unlike plasmid DNA degradation by human HD plasma which was inhibited by G-actin, NET degradation was insensitive to G-actin inhibition (**Fig. 2A and 2B**). These results suggested that while plasmid DNA was degraded by DNase I, NET degradation is mediated by other factors plasma factors. We measured DNase activity at 2.5%, 5% and 10% plasma dilutions (**Fig. S2C**) and fitted the data to curves to estimate the dilution required to yield 50% activity (D_50_) (**Fig. S2D**). Using a D_50_ cut-off of 15 we established that approximately 30% of the SARS-CoV-2 patient samples and 60% of microbial sepsis plasma samples exhibited significantly reduced DNase activity, compared to 10% of samples in the healthy donor group (**Fig. 2C**). Proteomic analysis uncovered significantly higher levels of plasma actin in SP and CP samples with low DNase activity suggesting that elevated actin levels in these patients were associated with lower DNase activity (**Fig. 2D**).

**Figure 2.**
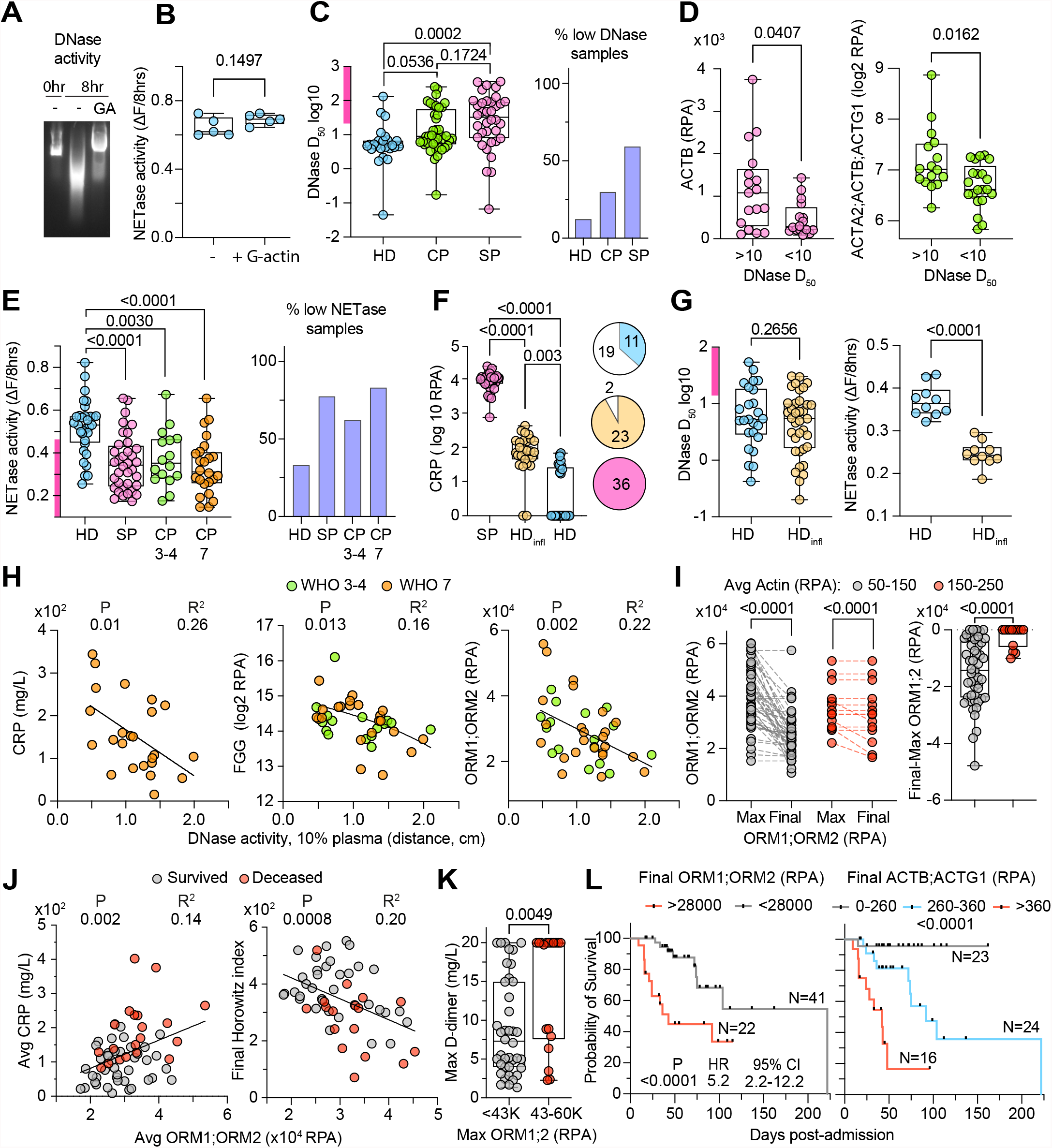
Low DNA and NET clearance activities in the plasma of healthy donors and patients with microbial sepsis and severe COVID-19 pneumonia. **A** and **B**. Plasmid DNA (A) and NET (B) degradation by HD plasma alone or in the presence of G-actin. **C**. Plasmid DNA degradation by 30 healthy donor (HD), 40 COVID-19 pneumonia (CP) and 36 microbial sepsis (SP) plasmas measured as D_50_ (% plasma required for 50% degradation) from raw data in Fig. S2C and S2D. Low activity range depicted by the purple bar on the y axis. Distribution of high and low DNase activity samples (right). **D**. Actin levels in SP and CP plasmas measured by MS proteomics and clustered by the corresponding DNase activity levels. Protein levels plotted as relative protein abundance (RPA). **E**. NETase activity in 30 HD, 43 CP and 36 SP plasma samples. Distribution of high and low NETase activity samples (right). **F**. CRP levels in HD, HD_infl_and SP plasma proteomes. **G**. DNase and NETase activity in HD and HD_infl_ plasmas. **H**. Correlation between raw DNase activity measurements at 10% plasma dilution and CRP, FGG or ORM proteins in WHO-7 and WHO-3-4 CP samples. **I**-**L**. 63 SARS-CoV-2 patients who reached a maximum WHO severity grade 7: **I**. Maximum and final ORM1;ORM2 values (left) and their difference (right) in relation to the longitudinal average actin values above (red circles) or below 150 RPA (grey circles) per patient. **J**. Correlation between the average ORM protein and clinical CRP levels. **K**. Maximum D-dimer readings in patients with maximum ORM1:ORM2 values above or below 42000 RPA. **L**. Probability of survival in patients with either high or low final ORM or actin protein levels. For Actin>360 or 260-360, hazard ratios (HR) =4.5 and 18.2, and 95% confidence interval (CI)= 1.3-15.8 and 4.9-67 respectively. Black bars indicate censoring events. Statistical analysis by Man-Whitney test for single comparisons, one-way Anova for multiple comparisons, simple linear regression for correlations, where P values appear on the left and R2 values on the right and Mantel-Cox survival analysis.

Low NETase activity afflicted approximately two-thirds of the samples we measured in both SP and CP cohorts (**Fig. 2E**). Surprisingly, we also detected comparable low NETase activity in 33% of the HD samples. To understand the differences within the HD pool, we analysed the plasma proteomes in two distinct HD cohorts from participants in the Avon Longitudinal Study of Parents and Children (ALSPAC) study^32, 33, 34^ segregated by low (HD) and high plasma levels of the novel GlycA inflammatory marker (HD_infl_)^35^. Many ALSPAC study participants have a high body mass index (BMI) and are insulin-resistant and hypertriglyceridemic. Consistent with our first HD cohort, we detected elevated C-reactive protein (CRP) levels in one third of HD samples (11/30), whereas nearly all HD_infl_ participants contained high CRP levels (28/30) (**Fig. 2F**). Remarkably, the two groups had comparable DNase activities, but NETase activity was significantly lower in all tested HD_infl_ samples (**Fig. 2G and S2D**). Hence, DNA clearance capacity is variable amongst healthy individuals and low NETase activity is associated with chronic inflammation.

Consistent with the ability of DNases to clear pro-inflammatory extracellular chromatin^19^, low DNase activity correlated with elevated CRP in WHO severity grade 7 CP plasmas and elevated FGG and complement factors that are linked to coagulation in all samples (**Fig. 2H and S3A**). Moreover, orosomucoid 1 and 2 (ORM1;OMR2) peptides were elevated in microbial sepsis and correlated with low DNase activity in CP samples independently of WHO severity grade (**Fig. 2H and S3A)**. High levels of ORM proteins are associated with high mortality in sepsis and hinted that low DNA degradation capacity in CPs was detrimental^36, 37, 38^. Low NETase activity correlated with elevated levels of complement cascade factors (C1QA, C1QB, C1QC) and apolipoproteins such as APOB and APOE (**Fig. S4**).

The association between DNase activity and ORM1;ORM2 protein levels independently of the WHO severity grade led us to employ ORM1;ORM2 as a surrogate marker to track DNase activity in a longitudinal set of 467 samples from 63 SARS-CoV-2 infected patients who reached maximum WHO severity grade 7 during hospitalization. We separated patients based on the survival outcome and plotted the cumulative daily average values from the readings of all patients (**Fig. S5A**). In most patients, average ORM1;ORM2 levels were the highest upon admission and decreased steadily thereafter in survivors, whereas they remained persistently high for up to 30 days post-admission in patient who did not survive. Actin levels began to decrease 20 days after hospital admission in survivors. In contrast, high actin levels persisted in patients that succumbed to the infection alongside a declining cumulative daily average Horowitz index (P/F ratio) (HI) and higher CRP levels between days 20-30. HI in patients that survived improved steadily from a daily average of 220 to above 400, whereas the group of deceased patients exhibited declining HI measurements over time. Moreover, patients who experienced thromboembolisms maintained high cumulative daily average FGG levels (**Fig. S5B**). These dynamics were reflected by the differences in the most proximal value to day 20 (Day20+/-), and the maximum and the average ORM and actin values per patient for the entire time period (**Fig. S5C and S5D**). Moreover, actin levels overwhelmingly decreased over time from their maximum levels in surviving patients, whereas actin levels had an upward trend over time in most non-survivors (**Fig. S5E**).

Consistent with the dependence of DNase activity on plasma actin levels, ORM protein levels in plasma remained high in most patients with high average actin levels (**Fig. 2I**). Moreover, the average ORM and actin protein levels positively correlated with average CRP levels per patient (**Fig. 2J and S6A**). Patients with elevated average ORM1;ORM2 levels exhibited poor lung function recovery as indicated by the final HI values (**Fig. 2J**) and lower difference between minimum and final values (**Fig. SB**). Furthermore, patients with high maximum ORM levels had significantly higher maximum D-Dimer readings, denoting the link between low DNase activity and coagulopathy (**Fig. 2K**). Consistent with these clinical measurements, patients with an average ORM1;ORM2 reading above 30000 RPA or elevated final readings experienced significantly higher rates of mortality (**Fig. 2L and S6C**). Given that high mortality rates occurred approximately 20-30 days post-admission, we also examined mortality in relation to ORM1;ORM2 readings from samples most proximal to day 20 post-admission. Patients that succumbed to the infection exhibited significantly higher ORM plasma levels during this critical time period and elevated mortality (**Fig. S5C and S6C**). Mortality also depended on plasma actin levels when patients were segregated based on their average, final or D20+/- actin levels (**Fig. 2L and S6C**). These findings applied to patients irregardless of age and co-morbidities since patients with different ORM protein level ranges had similar age ranges and rates of cardiovascular co-morbidities (**Fig. S6D**). Hence, low DNase activity was associated with higher actin levels and increased mortality in SARS-CoV-2 infected patients.

Given that plasma proteomics allows for the association of activities with multiple factors, we undertook a second gating approach to assign each patient sample to different DNase and NETase activity categories based on multi-factor proteomic profiling. Using this digital approach, we subsequently analysed the sample content per patient, rather than average levels of a single plasma marker. In contrast to the ORM-based analysis that examined all available samples per patient independently of the corresponding sample severity, this second approach employed only WHO severity grade 7 rated samples, restricting the analysis to 300 WHO-7 rated samples from 52 severe CP patients.

For DNase activity, we observed that measurements at 2.5% plasma segregated the 24 WHO-7 measured samples into high and low activity clusters (**Fig. 3A**). Separating these samples by survival outcome indicated that under these conditions, DNase activity could be used to predict mortality (P=0.023) with an area under the curve (AUC) value of 0.76 (**Fig. 3B and 3C**). Plotting the DNase measurements in 2.5% and 5% plasma dilutions or against the D_50_ indicated three categories of low, medium and high DNase samples (**Fig. S7A**). We classified into medium activity category, samples that were low by 2.5% plasma measurements but exhibited higher activity in 5% and 10% plasma measurements. We identified mannose binding lectin-2 (MBL2), complement factor H related 1 (CFHR1) and apolipoprotein C1 (APOC1) as markers that could segregate samples into different DNase activity groups (**Fig. 78B-D**). MBL2 was particularly interesting as it was present at significantly lower levels in all WHO-7 samples from the deceased group (**Fig. S7E**). A multi-gating approach that was based on CHFR1/APOC1/MBL2/ and FGB/C1QA combinations allowed us to assign all 300 WHO-7 samples into low, medium and high DNase activity groups (**Fig. S7F**). Using these assignments, we examined the distribution of samples in the surviving and deceased groups which indicated that low and medium DNase activity samples were predominately found to be associated with non-survivors (**Fig 3E and 3F**).

**Figure 3.**
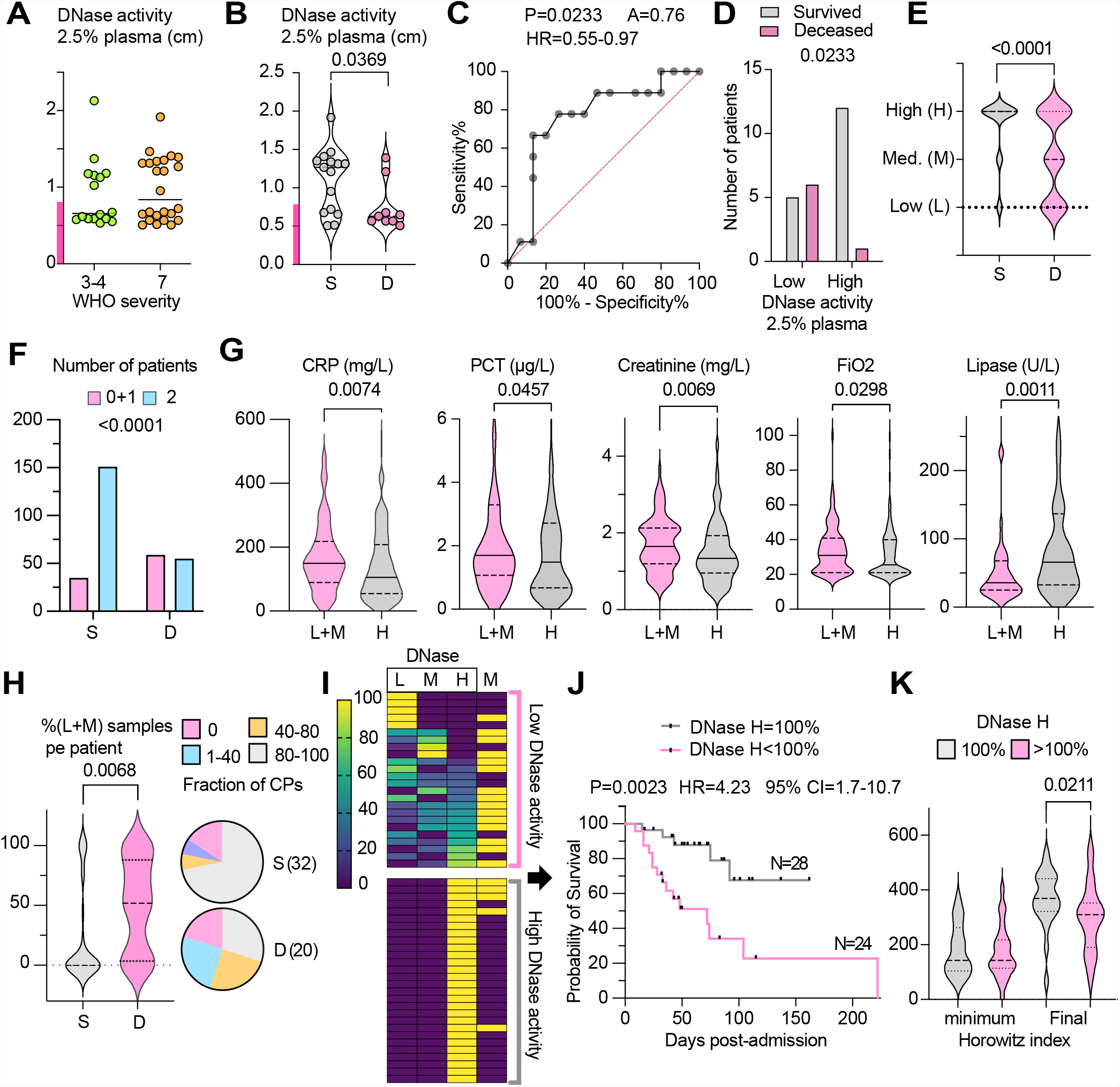
Low DNase activity profiles are associated with high mortality in patients with severe SARS-CoV-2 infection. **A**. DNAse activity in 40 CP plasmas measured at 2.5% plasma dilution segregated by WHO severity grade. Purple bar indicates the low activity range. **B**. DNAse activity in 25 WHO-7 CP plasmas measured at 2.5% plasma dilution and segregated by whether patients survived (S) or died (D). **C** and **D**. AUC of patients and distribution of surviving and deceased patients by low and high DNase activity in (B). **E**. Distribution of 300 WHO-7 samples from 52 CP patients with maximum WHO severity grade 7, based on low (L), medium (M) and high (H) DNase activity assigned by proteomic profiling in surviving or decease patients. **F**. Number of low and medium (L+M) or high (H) DNase activity samples in surviving or decease patients. **G**. Clinical measurements that are significantly different in the 300 WHO-7 CP samples segregated by profiled DNase activity into low and medium or high activity groups. **H**. Violin plot of the fraction of low+medium DNase activity samples per patient segregated by disease outcome and pie chart of the fraction of patients in the two groups that fall within different sample content ranges. **I**. Heat map of 52 max-WHO-7 patients ranked by their content of assigned DNase activity samples. Rows depict individual patients. The first three columns depict the percentage of low, medium and high assigned DNase activity samples and the fourth column mortality (M) where yellow indicates deceased patients. **J**. Survival analysis of CP patients containing low+medium DNase activity samples (DNase H<100%, pink) or a group that contains 100% high DNase activity samples (grey). Black bars indicate censoring events. **K**. Comparison of minimum and final Horowitz index values per patient segregated by the fraction of profiled high DNase activity (2) samples. Statistical analysis by Man-Whitney test for single comparisons, Mantel-Cox survival analysis, Fisher’s exact test for distribution and AUC using the Wilson/Brown method.

Next, we examined whether the low, medium and high DNase activity samples were associated with differences in over 60 clinical parameters. Low and medium activity samples were indistinguishable by most clinical parameters. Therefore, we decided to present them as a single low+medium activity group. Low-medium DNase activity samples exhibited higher CRP, procalcitonin (PCT) and creatinine levels and lower lipase levels, indicative of elevated inflammation, bacterial co-infection, and kidney and pancreatic damage (**Fig. 3G**). Moreover, these samples were associated with significantly higher oxygen requirements (FiO_2_) (**Fig. 3G**). We also evaluated the distribution of samples in each patient and found that most survivors either lacked or had only a small fraction of low-medium DNase activity samples compared to the group of deceased patients (**Fig. 3H**). Clustering of patients based on their DNase activity sample content indicated that deaths occurred predominately in patients with some degree of low DNase activity (**Fig. 3I**). Patients that contained low and medium DNase activity samples exhibited a 4.2-fold increase in mortality and lower lung function recovery compared to patients that contained only high DNase activity samples (**Fig. 3J and 3K**). Hence, DNase activity assignments base on proteomic profiling, identified high-risk patients that were associated with increased pathology and mortality within the severe patient cohort.

Next, we classified samples into NETase activity categories by employing 3 plasma proteins that correlated with NETase activity in the 24 WHO-7 samples. The neutrophil chemokine pro-platelet basic protein (PPBP, CXCL7) correlated with NETase activity across WHO severity grades, alongside complement component 4 binding protein beta (C4BPB) and the protease inhibitor alpha-2-macroglobulin (A2M) (**Fig. S8A**). There was a significance difference in the levels of these factors in the 300 longitudinal samples from survivors and non-survivors consistent with a trend for lower NETase activity in non-survivors (**Fig. S8B**). Low NETase activity samples were associated with a PPBP^low^;A2M^high^ proteomic profile (**Fig. S8C**) whereas high NETase activity samples were associated with high C4BPB levels (**Fig. S8A**). Interestingly, plotting all 300 samples on a PPBP/A2M graph indicated a higher prevalence of PPBP^low^;A2M^high^ samples in the deceased group that were absent in survivors (**Fig. S8D**). Consistently, the samples that fitted to a low NETase activity profile were associated predominately with non-survivors (**Fig. 4A and 4B**). A combined DNase-NETase classification into 6 groups indicated that survivors contained predominately a combination of high DNase activity and medium and high NETase activity samples, whereas non-survivors contained a high proportion of low DNase and NETase activity samples (**Fig. 4C and 4D**). Notably, profiled DNase and NETase activities clustered independently, consistent with the notion that they are driven by different pathways.

**Figure 4.**
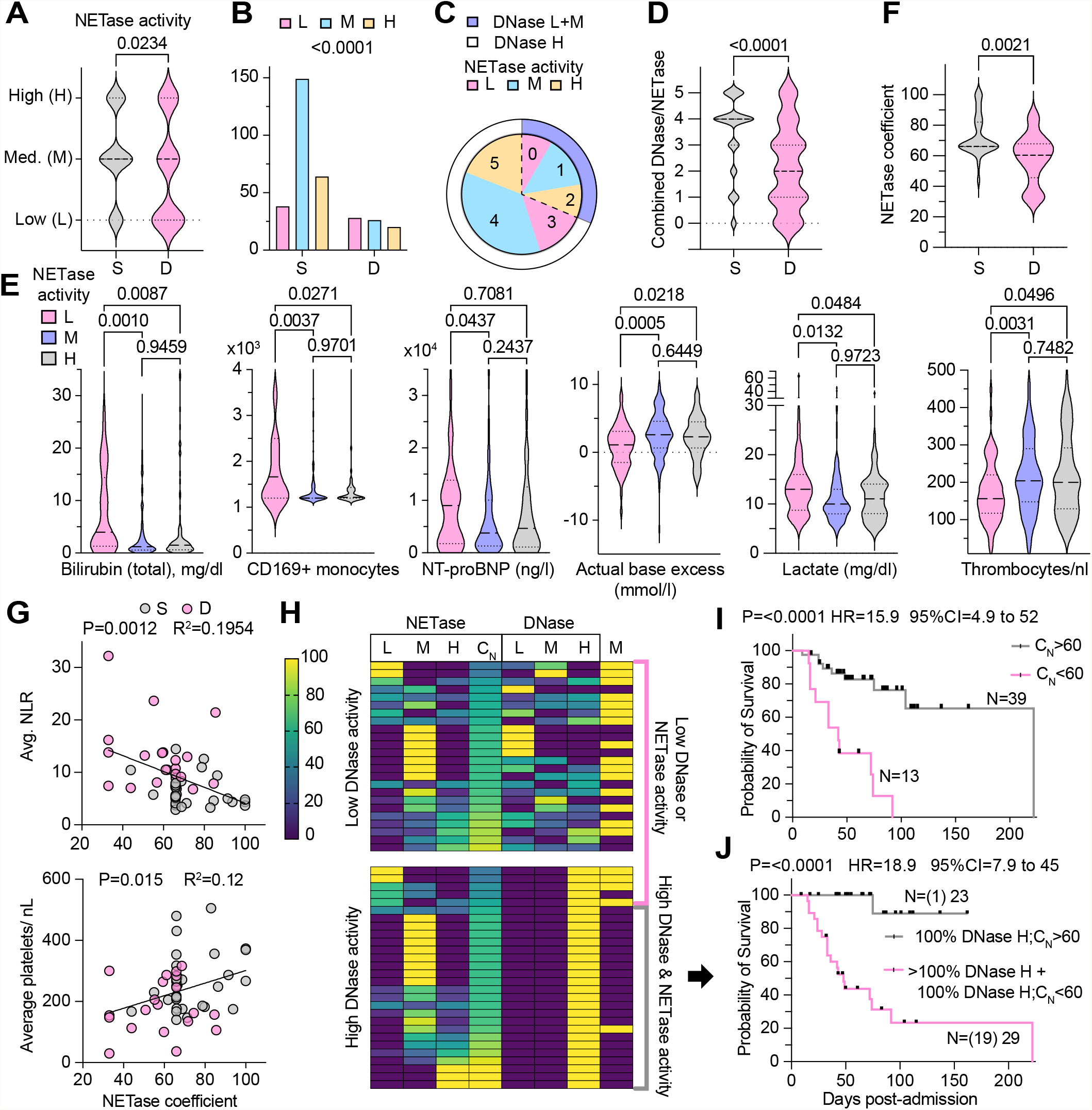
Low NETase activity profiles are associated with high mortality in patients with severe SARS-CoV-2 infection. **A**. Distribution of 300 WHO-7 samples from 52 CPs who either survived (S) or died (D), based on profiled low (L), medium (M) and high (H) NETase activity from plasma proteome profiles. **B**. Number of profiled low, medium and high NETase activity samples from surviving or deceased patients. **E**. AUC plot of data in (A). **D**. Distribution of samples in surviving and deceased patients based on the combined profiled DNase and NETase activity assignments. The pie chart depicts the fraction of samples in these categories. The inner circle depicts profiled NETase and the outer ring DNase activity assignments. **E**. Clinical measurements that are significantly different in the 300 WHO-7 CP samples segregated by profiled NETase activity. **F**. NETase coefficient (C_N_) per patient in patients that survived or died, calculated as 0.33*(%L samples)+0.66*(%M samples)+1*(%H samples). **G**. Correlations between the C_N_ and the average number of platelets or the neutrophil to lymphocyte ratio (NLR) in all samples per patient including lower WHO severity. **H**. Heat map of 52 max WHO-7 CP patients by whether the exhibited a 100% high DNase activity content and ranked by their C_N_. Each rows shows an individual patient. Columns depict the percentage of assigned low, medium and high NETase and DNase activity samples per patient, the C_N_ and mortality (M) where yellow denotes deceased patients. **I**. Survival analysis of patients in (H) with a NETase coefficient either above (grey) or below 60 (pink). **J**. Survival of patients with either a combined high DNase and NETase activity (C_N_ above 60 and 100% high DNase activity sample content, grey) or low DNase or NEtase activity (high DNase activity sample content below 100% or C_N_ below 60, pink), as shown by brackets in (H). Black bars indicate censoring events. Statistical analysis by Man-Whitney test for single comparisons, one-way Anova for multiple comparisons, Mantel-Cox survival analysis, Fisher’s exact test for contingency analysis and simple linear regression for fitting.

Next, we explored how clinical parameters varied in samples with different profiled NETase activities. Bilirubin and pro-BNP were significantly elevated in low and medium NETase activity compared to high NETase activity samples, indicating increased red blood cell lysis and signs of heart failure (**Fig. 4E**). Strikingly, the number of circulating activated CD169+ macrophages was specifically increased in low NETase activity samples. Higher lactate levels and a base deficit associated with acidosis were also suggestive of poor lung function with a metabolic impact in low NETase activity samples. Furthermore, the increased thrombocytopenia in these samples was a strong indication of coagulopathy.

To obtain an overview of sample distribution we employed a formula to calculate a NETase coefficient (C_N_) for each patient as the weighted aggregate of NETase activity sample distribution. Survivors exhibited higher NETase coefficients than the deceased patients which positively corelated with the average number of platelets and neutrophil to lymphocyte ratio calculated from all available samples per patient (**Fig. 4F and 4G**). Ranking of patients according to their C_N_ and DNase high activity content indicated high mortality rates in patients with a significant content of low NETase activity samples (**Fig. 4H, left**). Interestingly, 22 patients contained samples that were classified exclusively to one NETase class and 30 patients displayed a mixed assortment, suggesting NETase activity fluctuations in some patients (**Fig. 4H, right**). Patients with a NETase coefficient below 60 exhibited a 15-fold higher mortality rates than patients with a coefficient above 60 (**Fig. 4I**). Given the lack of overlap between DNase and NETase activity distributions in patients, we examined the combined effect of both activities on survival. Remarkably, only 1 patient died amongst the 23 patients that had a C_N_ above 60 and all of their samples classified as high DNase activity as opposed to 19 deaths in the 29 patients that contained low DNase activity samples or a combination of high DNase activity and low NETase (C_N_<60) samples (**Fig. 4H and 4J**). Assuming that all censored patients survived after being discharged, the hazard ratio jumped from 18.9 to 22.8 suggesting that a combination of both non-redundant activities protects against mortality in critically ill patients (**Fig. S8E**).

To further validate the DNase and NETase activity assignments, we searched for factors that were significantly different amongst the combined DNase/NETase classified samples. Several of these proteins such the apolipoproteins APOA1, APOB, APOC1, APOL1, as well as immunoglobulin heavy constant gamma 4 (IGHG4) also correlated with direct DNase and NETase activity measurements, (**Fig. 5A**). We also observed consistent changes in these proteins as well as A2M and MBL2 in the murine fungal sepsis model after treatment with DNase I or anti-histone antibodies which supported a functional link between extracellular chromatin clearance and the levels of these proteins (**Fig. 5B**). Both treatments blocked hypothermia, T cell death in the spleen and actin release (**Fig. 5B-E**), suggesting a negative feedback mechanism where the clearance of extracellular chromatin by DNases inhibits actin-release. Hence, such therapies could be beneficial for patients by limiting T cell death and actin release in the circulation.

**Figure 5.**
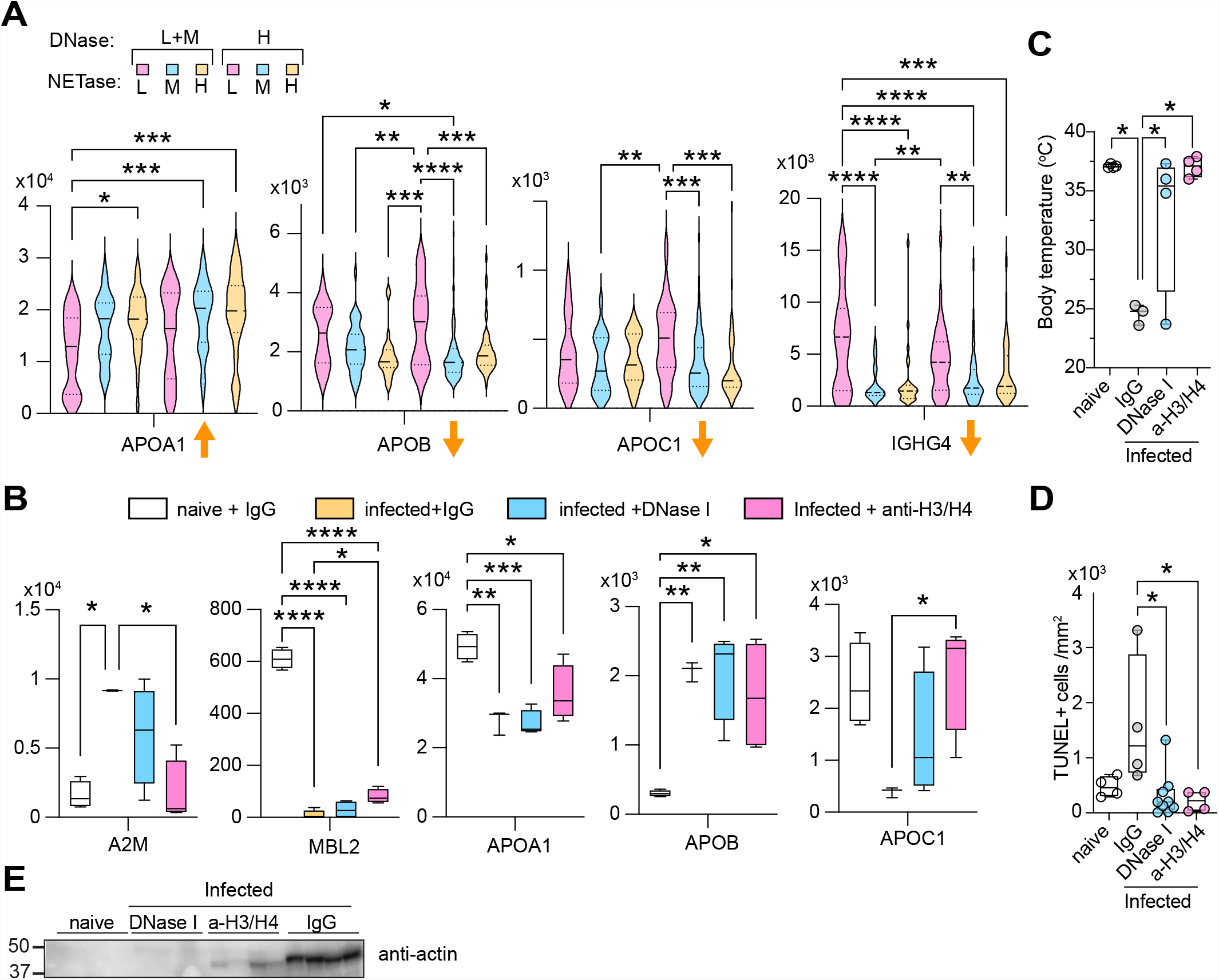
Validation of plasma factors correlating with DNase and NETase activity profiles in SARS-CoV-2 patients and the murine sepsis model. **A**. Relative protein levels (RPA) for selected proteins that in 300 WHO-7 samples that change significantly when samples are segregated by combined profiled DNase and NETase activity assignments. Arrows indicated whether these proteins positively or negatively correlate directly with NETase measurements in WHO-7 CP samples in Fig S5. **B-E**. Selected plasma proteins (B), body temperature (C) number of TUNEL+ T cells in the spleens (d) and western immunoblotting for plasma actin (e) from naive WT mice or infected with *C. albicans* and treated with control IgG or anti-histone H3 and H4 antibodies or DNase I. Statistical analysis by one-way Anova for multiple comparisons, where * 0.05-0.01, ** 0.01-0.001, *** 0.001-0.0001 and **** <0.0001.

Our findings uncover a critical role for extracellular DNA clearance pathways in the survival of patients with severe SARS-CoV-2 infection, validating the pathogenic role of extracellular chromatin. We demonstrate that defects in these pathways are prevalent in patients with acute infections and deficiencies can be pre-existing or exacerbated by the release of actin. Surprisingly, NET clearance appears to involve different non-redundant mechanisms than the pathways degrading naked DNA. Although both assays were able to identify critical samples linked to patient survival, the activity patterns did not overlap, and these activities behaved differently with respect to G-actin inhibition. Several proteins that correlated with these activities may play functional roles or denote downstream consequences of variations in DNA clearance such as the complement and coagulation factors which are consistent with the link between the complement cascade, NET formation and thrombosis^3, 39, 40, 41^. The correlations and functional association with apolipoprotein levels suggest a link between DNA clearance and lipid metabolism. Furthermore, we report for the first time, variations in DNase and NETase activity in healthy individuals which may pre-dispose them to severe pathology upon infection. While actin and hyper-inflammation may promote such deficiencies it is likely that many patients exhibit low DNA clearance capacity prior to infection. The link between chronic inflammation in heathy individuals and low NETase activity provides one explanation for the association of severe COVID-19 pneumonia with obesity and cardiovascular conditions. However, we did not observe differences in NETase and DNase activity distribution that could be associated with cardiovascular co-morbidities suggesting that deficiencies may be determined pre-clinically or triggered during infection (**Fig. S9A**). The impact of low DNA clearance capacity on mortality suggests that therapies that target extracellular chromatin may prove beneficial. Functional proteomic profiling allows the identification of pathologically relevant markers in samples that are otherwise difficult to distinguish by broad analysis (**Fig. S9B**). Furthermore, it enables multiple activity tracing in large sample sets and the identification of high-risk individuals within cohorts of patients with severe disease symptoms that may not be easily distinguishable by conventional clinical parameters.

## Data Availability

All data produced in the present study are available upon reasonable request to the authors

## Acknowledgements

We are extremely grateful to the patients and their families. We thank Andrew Aswani for sourcing sepsis patient samples. This research was funded in whole, or in part, by the Wellcome Trust (FC0010129, FC001134). For the purpose of Open Access, the author has applied a CC BY public copyright licence to any Author Accepted Manuscript version arising from this submission. This work was supported by the Francis Crick Institute which receives its core funding from the UK Medical Research Council (FC0010129, FC001134), Cancer Research UK (FC0010129, FC001134) and the Wellcome Trust (FC0010129, FC001134). I.V.A was funded by an EMBO LTF (ALTF 113-2019). S.T.C. was funded by BHF project grant PG/18/45/33814. We also thank all patients and their families as well as the Charite PA-COVID-19 study group: Sebastian Albus, Florian Alius, Tim Andermann, Stefan Angermair, Uwe D. Behrens, Laure Bosquillon de Jarcy, Markus Brack, Astrid Breitbart, Felix Bremer, Felix Bremer, Dana Briesemeister, Victor M. Corman, Thomas Cronen, Chantip Dang-Heine, Sophy Denker, Phillip van Dijck, Jan-Moritz Doehn, Christian Drosten, Kai-Uwe Eckardt, Marius A. Eckart, Andreas Edel, Lucas Elbert, Matthias Endres, Philipp Enghard, Matthias Felten, Carolin Ferse, Simon Fraumann, Nikolaj Frost, Carmen Garcia, Dominik Geus, Gisele J. Godzick-Njomgang, Daniel Grund, Sascha S. Haenel, Jens K. Haumesser, Julia Heeschen, Friederike L. Hefele, Katrin M. Heim, Anne Herholz, Vera Hermanns, Moritz Hilbrandt, David Hillus, Stefan Hippenstiel, Andreas Hocke, Johannes Hodes, Ralf-Harto Hübner, Michael Hummel, Till Jacobi, Linda Jürgens, Christof von Kalle, Ute Kellermann, Ye-Ji Kim, Malte Kleinschmidt, Philipp Knape, Samuel Knauß, Roland Körner, Alexander Krannich, Philipp M. Krause, Lucie Kretzler, Jan M. Kruse, Liron Lechtenberg, Lukas Lehner, Yaosi Li, Linna Li, Emma Lieker, Tilman Lingscheid, Felix Machleidt, Elena Madlung, Josef Mang, Luise Martin, Nikolai Menner, Lil Meyer-Arndt, Agata Mikolajewska, Belén Millet Pascual-Leone, Mirja Mittermaier, Martin Möckel, Luisa Mrziglod, Nadine Muller, Holger Müller-Redetzky, Sergej Münzenberg, Christopher Neumann, Michaela Niebank, Nadine Olk, Bastian Opitz, Eva Pappe, Panagiotis Pergantis, Frieder Pfäfflin, Lennart Pfannkuch, Moritz Pfeiffer, Thuy N. Pham, Peter Radünzel, Matthias Raspe, Nicola Reck, Johannes Rein, Jana Riecke, Teresa Ritter, Anne Ritter, Maria Rönnefarth, Christoph Ruwwe-Glösenkamp, Jacopo Saccomanno, Leif-Erik Sander, Birgit Sawitzki, Laura K. Schmalbrock, Sein Schmidt, Solveig Schönberger, Rosa C. Schuhmacher, Dirk Schürmann, Mariana Schürmann, Georg Schwanitz, Dominik Soll, Claudia Spies, Miriam S. Stegemann, Fridolin Steinbeis, Britta Stier, Paula Stubbemann, Norbert Suttorp, Christoph Tabeling, Frank Tacke, Bettina Temmesfeld-Wollbrück, Charlotte Thibeault, Pinkus Tober-Lau, Sascha Treskatsch, Denise Treue, Alexander Uhrig, Markus Vogtmann, Nadia A. de Vries, Lisa-Marie Wackernagel, Susanne Weber, Steffen Weber-Carstens, Anne Wetzel, Isabelle Wirsching, Marcel Wittenberg, Martin Witzenrath, Christian Wollboldt, Alexander Wree, Yinan Wu, Berna Yedikat, Tamar Zhamurashvili, Daniel Zickler, Christian Zobel, Thomas Zoller, Saskia Zvorc and Selina Greuel.

## Author Contributions

I.V.A, D.H., M.I., M.I.T. and C.F.M.T. designed and performed experiments and analysis, S.P, V.D and M.W. performed mass spectrometry measurements and analysis, E.T.H., J.I, S.G., H.R, L.L., E.T.H. and F.K. supervised the SARS-CoV-2 patient study, S.D. K.S. and C.B. supervised the microbial sepsis patient study, S.T.C. and J.E.D. provided healthy donor samples, M. R. directed the proteomic analysis. V.P directed the study, performed analysis and wrote the manuscript.

## Materials and Methods

### Human healthy donor and clinical samples of Sepsis and COVID-19 patients

For the in vitro neutrophil experiments, peripheral blood was isolated from consenting healthy adult volunteers, according to approved protocols of the ethics board of the Francis Crick Institute and the Human Tissue act. Sepsis patient samples were provided by the Hannover Medical School approved the ethics committee under the study protocol (No. 2786-2015). Written informed consent was obtained from participants or authorized representatives. The study was performed in accordance with the ethical standards laid down in the 1964 Declaration of Helsinki and its later amendments. No funding specific to this project was received. Sampling was performed as part of the Pa-COVID-19 study, a prospective observational cohort study assessing pathophysiology and clinical characteristics of patients with COVID-19 at Charité Universitätsmedizin Berlin^42^. All patients with SARS-CoV-2 infection proven by positive PCR from respiratory specimens and willing to provide written informed consent are eligible for inclusion. Exclusion criteria are refusal to participate in the clinical study by patient or legal representative or clinical conditions that do not allow for blood sampling. The study assesses epidemiological and demographic parameters, medical history, clinical course, morbidity and quality of life during hospital stay of COVID-19 patients. Moreover, serial high-quality bio-sampling consisting of various sample types with deep molecular, immunological and virological phenotyping is performed. Treatment and medical interventions follow standard of care as recommended by current international and German guidelines for COVID-19. Severity of illness in the present study follows the WHO ordinal outcome scale. The Pa-COVID-19 study is carried out according to the Declaration of Helsinki and the principles of Good Clinical Practice (ICH 1996) where applicable and was approved by the ethics committee of Charité-Universitätsmedizin Berlin (EA2/066/20). Plasmas of healthy donors with and without inflammation signatures were selected from participants in the Avon Longitudinal Study of Parents and Children (ALSPAC)^32, 33, 34^. All ethical approvals conformed to the Declaration of Helsinki and all biological samples used in the study were collected in accordance with the Human Tissue Act (2004).

### Mouse strains and ethics

All mice were bred and maintained in a pathogen free, 12-hour light-dark cycle environment. All experiments were conducted with age-matched and cage-controlled, 8 to 12-week-old female WT C57BL/6J and TCRα-/- (*Tcra*^*tm1Phi*^) mice, according to local guidelines and UK Home Office regulations under the Animals Scientific Procedures Act 1986 (ASPA).

### Sepsis *in vivo* model

Wild-type Candida albicans (*C. albicans*, clinical isolate SC5314) was cultured overnight in yeast extract peptone dextrose (YEPD; Sigma) medium, while shaking at 190RPM at 37°C. On the subsequent day, overnight culture was sub-cultured in YEPD and allowed to grow for approximately 4 hours, until an optical density (A600) of 0.4-0.8 was measured. Subcultures were microscopically examined for lack of hyphae, washed in sterile phosphate-buffered saline (PBS) and spun for 10min at 1000xg at RT. Mice were intravenously injected with 5×10^5^ yeast per mouse via the tail vein. The weight and rectal temperature of the mice were recorded daily over the course of infection to track health status. A body temperature below 32°C, a weight loss superior to 80% of initial weight accompanied by slow movement and non-responsiveness were considered collectively as septic shock and the humane endpoint for the mice. The mice were culled via cervical dislocation or by lethal dose of pentobarbital (600mg/kg) with mepivacaine hydrochloride (20mg/ml).

For degradation of circulating DNA *in vivo*, mice were treated with deoxyribonuclease I (DNase I) from bovine pancreas (Sigma; 2000 units/mouse) daily via intraperitoneal injection, starting on the day prior to infection (D-1). For *in vivo* histone neutralisation, the mice received combined anti-histone 3 (Merck Millipore; 07-690) and anti-histone 4 antibodies (Merck Millipore; 04-858) or control polyclonal rabbit IgG (BioXCell). Anti-histone antibodies were dialysed and injected intraperitoneally, starting on D-1 (200μg/mouse) and daily afterwards (200μg H3 and 100μg H4). Treatments were given daily until completion of the experiment.

### Histology and immunofluorescence imaging

For mouse tissue staining, freshly extracted spleens were directly embedded in optimal cutting temperature (OCT) compound cryo-embedding media (VWR Chemicals BDH) and frozen. Sections were cut to 8μm thickness on positively charged glass slides. To prepare samples for staining, the glass slides were air dried for 20min at RT, washed in PBS and fixed for 10min with 4% paraformaldehyde (PFA; Sigma) at RT.

For staining of paraffinized and cut COVID patient spleens, slides were first baked at 60°C for 1hr. To remove paraffin, samples received 3 sequential baths of 5mins in Neo Clear and were then rehydrated with sequential 100%, 96%, 80%, 70% and 50% EtOH baths, 5min each, and washed. Antigen retrieval was performed with Dako Target Retrieval Solution pH9, for 45min at 97°C.

All tissues were permeabilized with 0.5% Triton X-100 in PBS for 20min at RT. For terminal deoxynucleotidyl transferase dUTP nick end labeling (TUNEL), the Click-iT TUNEL Alexa Fluor 594 Imaging Assay kit (Invitrogen) was used and instructions from the manufacturer were followed. After TUNEL staining, non-specific binding was blocked with 2% BSA (Sigma) and 2% donkey serum (Sigma) in PBS for 1hr at RT. Samples were then stained overnight in a dark humified chamber with the following primary antibodies in blocking buffer: anti-mouseCD3 (BioLegend; clone 17A2), anti-CD3 (Abcam; ab5690), anti-CD169 (BioLegend; clone 3D6.112) and anti-MPO (R&D Systems; AF3667). For secondary staining, tissues were stained for 2hrs in a humidified dark chamber at RT with the following labelled secondary antibodies in blocking buffer: donkey anti-rabbit IgG (Invitrogen) and donkey anti-goat IgG (Invitrogen). Nuclear staining with 4’,6-diamidino-2-phenylindole dihydrochloride (DAPI; Invitrogen) was added with the secondary staining. All stained tissue sections were mounted in ProLong Gold (Molecular Probes). Images were taken using the Leica TCS SP5 inverted confocal microscope (20x, 40x, 63x original magnification) and analysis was performed using Fiji/ImageJ version 2.0.0 software.

### Neutrophil isolation from peripheral human blood

Peripheral venous blood collected in EDTA tubes was layered on Histopaque 1119 (Sigma-Aldrich) and centrifuged for 20 min at 800xg. The plasma was collected and centrifuged for a second time. The neutrophil layer was collected and washed in Hyclone Hank’s Balanced Salt Solution (HBSS) -Ca, -Mg, -Phenol red (GE Healthcare) supplemented with 10mM HEPES (Invitrogen) 0.1% plasma and further purified with a discontinuous Percoll (GE Healthcare) gradient consisting of layers with densities of 85%, 80%, 75%, 70% and 65% and centrifugated for 20 min at 800xg. Neutrophil enriched layer were collected and washed. Neutrophil purity was assessed via flow cytometry.

### Plasmid DNA degradation assay

1μg of a 6250 bp plasmid was incubated with 10 mM Tris, 2.5 mM MgCl_2_ and 2.5 mM CaCl_2_ at a pH 7.4 with 2.5/5 or 10 % plasma in a final volume of 20μl for 2h at 37°C. The sample was then run in 1% agarose gel electrophoresis for 30min. The images acquired from the gel were then analysis using Fiji. For the analysis; line profiles of each well were measured and the migrated distance of the DNA was quantified. The distance corresponding to the maximum intensity point (representing the most abundant fragment size) was plotted at the different plasma % for each plasma sample.

### Time-lapse imaging and NET degradation quantification

5×10^4^ human neutrophils were seeded in a black 96-well plate (PerkinElmer) in HyClone HBSS +Ca, +Mg, - Phenol red (GE Healthcare) containing 10mM HEPES (Invitrogen) and 3% plasma from healthy, septic or COVID-19 donors. We stained the DNA of life cells with 4ug/ml Hoechst (membrane permeable; Thermo Scientific) and of dead cells with 0.2μM Sytox-green (membrane impermeable; Invitrogen). The cells were imaged on an inverted Nikon wide-field microscope system at 37°C and CO_2_ (5%). Four fields of view were acquired per well every 10 mins for 10-15 hrs using a 40x objective.

Quantification of NET degradation was performed by automatically identifying individual NET events in four regions of interest per well and monitoring Sytox signal during the course of the acquisition. Sytox signal changes were measured for NETs formed in the first 400 min of the acquisition. The average and standard deviation of difference of signal between the maximum intensity point corresponding to an initial NET stage and the minimum intensity point corresponding to the NET signal at the end of the acquisition for each individual NET was plotted for each plasma sample.

### Western blot analysis

Samples were boiled in a sodium dodecyl sulphate (SDS) buffer containing dithiothreitol (DTT) and resolved by polyacrylamide gel electrophoresis (SDS-PAGE) on a Criterion TGX precast gel (Any-KD; Bio-Rad Laboratories). Proteins were then transferred to a PVDF membrane (Bio-Rad Laboratories) via semi-dry transfer. The membrane was blocked with 5% bovine serum albumin (BSA; Fisher Scientific) in Tris-buffered saline with 0.1% Tween 20 (TBS-T). DNase I was detected with anti-DNase I and HRP–conjugated goat anti-rabbit (Thermo Scientific). Fold increase or decrease of DNase I levels were compared to levels detected in plasma from healthy volunteers. H3 was detected with anti-histone 3 (Milipore) and HRP–conjugated goat anti-rabbit (Thermo Scientific). Actin was detected with anti-actin (Milipore) and HRP– conjugated donkey anti-mouse (Thermo Scientific) antibodies.

### DNA quantitation

Circulating DNA in plasma was quantified using the Quant-iTTM PicoGreen dsDNA assay kit (Thermo Fisher Scientific). The fluorescent signal (excitation at 488nm) was measured using a spectrophotometric microplate reader (Fluostar Omega, BMG labtech).

### Plasma sample preparation for proteomic analysis

Healthy donor and patient plasma samples were randomised and plated in a 96-well plate (Eppendorf). Protein/peptide extraction and proteomics analysis was performed following the protocol described in detail in Messner et al., 2020^24^. 5μL of plasma was denatured in 50μl 8M Urea (Honeywell Research Chemicals), 100mM ammonium bicarbonate (ABC, Honeywell Research Chemicals) and reduced with 5μL of 50mM dithiothreitol (DTT, Sigma Aldrich) at 30°C for 1 hour. Followed by alkylation with 5μL of 100mM iodoacetamide (IAA, Sigma Aldrich) at 23°C for 30 minutes in the dark. The samples were diluted with 340μL of 100mM ABC and 220μL was added to trypsin solution (Promega) for protein digestion at a trypsin/protein ratio of 1/40 and incubated at 37°C overnight (17h). Quenching of digestion was done by the addition of 25μL of 10% v/v formic acid (FA, Thermo Fisher Scientific). Rounds of solid phase extraction clean-up steps were performed with the use of C18 96-well plates (BioPureSPE Macro 96-Well, 100mg PROTO C18, The Nest Group) as described previously in Messner et al. 2020^24^. Methanol (Fisher Chemicals), 50% v/v acetonitrile (ACN, Fisher Chemicals) or 0.1% v/v FA was used at each centrifugation step as required. After final elution, the collected peptide material was dried by a vacuum concentrator (Eppendorf Concentrator Plus) and redissolved in 50μl 0.1% v/v FA, to be processed by liquid chromatography-mass spectrometry.

### Liquid chromatography-mass spectrometry

1μg of protein digest (peptides) was injected and analysed on a nanoAcquity Liquid Chromatograph (Waters) coupled to a TripleTOF 6600 Mass Spectrometer (Sciex) at a flow-rate of 5μl/min. This was followed by a separation using a Waters HSS T3 column (150mm x 300μm, 1.8μm particles) in 20-minute non-linear gradients starting with 3% B up to 40% B (Buffer A: 0.1% v/v FA; Buffer B: ACN / 0.1% v/v FA). A data independent acquisition (DIA/SWATH) method was used, with MS1 scan from m/z 400 to m/z 1250 and 50ms accumulation time followed by 40 MS2 scans of 35ms accumulation time with variable precursor isolation width covering the mass range from m/z 400 to m/z 1250. Ion source gas 1 (nebulizer gas), ion source gas 2 (heater gas) and curtain gas were set to 30, 15 and 25 respectively. The source temperature was set to 450°C and the ion spray voltage to 5500V. Injections of samples took place in a random order.

The “gas-phase fractionation” methodology was used to generate the library out of pooled peptide digest of all plasma samples with the use of a LC-MS/MS method as mentioned before. 3μg of protein digest was separated with a 60-minute linear gradient (3% B to 40% B). Injections were performed at the mass ranges of: 400 – 500 m/z, 495 – 600 m/z, 595 – 700 m/z, 695 – 800 m/z, 795 – 900 m/z, 895 – 1000 m/z, 995 – 1100 m/z, 1095 – 1250 m/z. The precursor selection windows were 2 m/z (1 m/z overlap). DIA-NN 1.7.10 proteomics analysis software^43^ was used for the library preparation with Mus musculus (mouse) UniProt (UniProt Consortium, 2019) isoform sequence database (UP000000589) to annotate the library. To quantify proteins, raw data acquired were processed with DIA-NN 1.7.10 with the “robust LC (high precision)” mode with MS2, MS1 and scan window size set to 20ppm, 12ppm and 8 respectively.

### Correlation and statistical analysis

All correlation and fitting analysis were performed using GraphPad Prism software aided by sorting and grouping samples using Microsoft Excel. Thresholds for identifying and grouping patients into groups were determined using frequency analysis in Excel. Single comparison statistical significance was assessed by an unpaired, two-tailed Student’s t-test. Mann-Whitney test for single comparisons, Fisher’s exact test for contingency, simple linear regression, or non-linear exponential or Gaussian distribution fitting. Statistical analysis was performed on GraphPad Prism software.

---

**Supplemental Figure 1.**
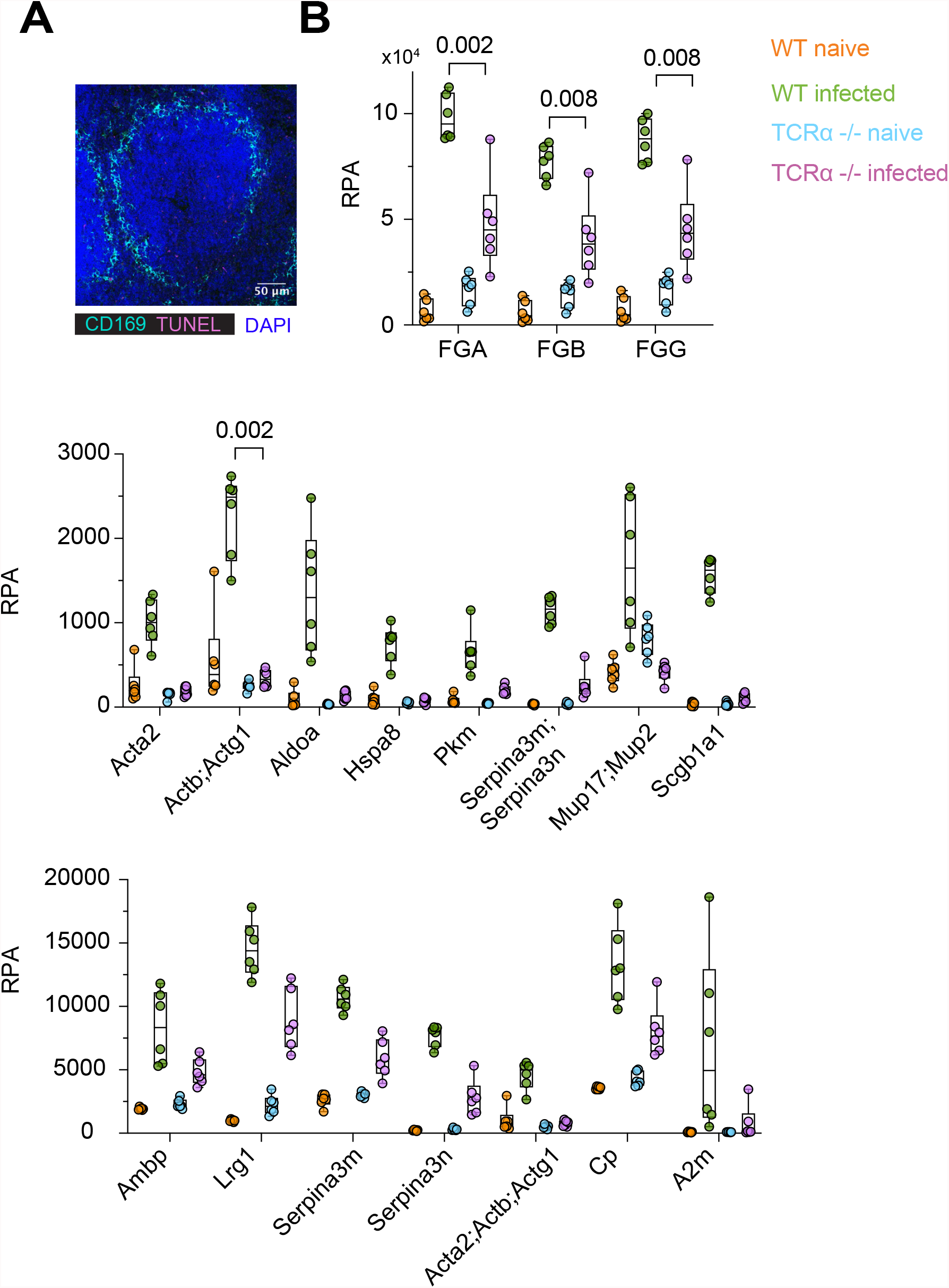
Systemic candidiasis triggers T cell-associated changes in plasma proteins. **A**. Representative immunofluorescence confocal microscopy micrograph of a naïve WT spleen stained for CD169, DAPI and TUNEL. **B**. Relative protein abundance (RPA) of selected proteins in the plasma from WT and TCRα-deficient mice, either naïve or infected intravenously with *C. albicans*. Statistical analysis by one-way Anova test.

**Supplemental Figure 2.**
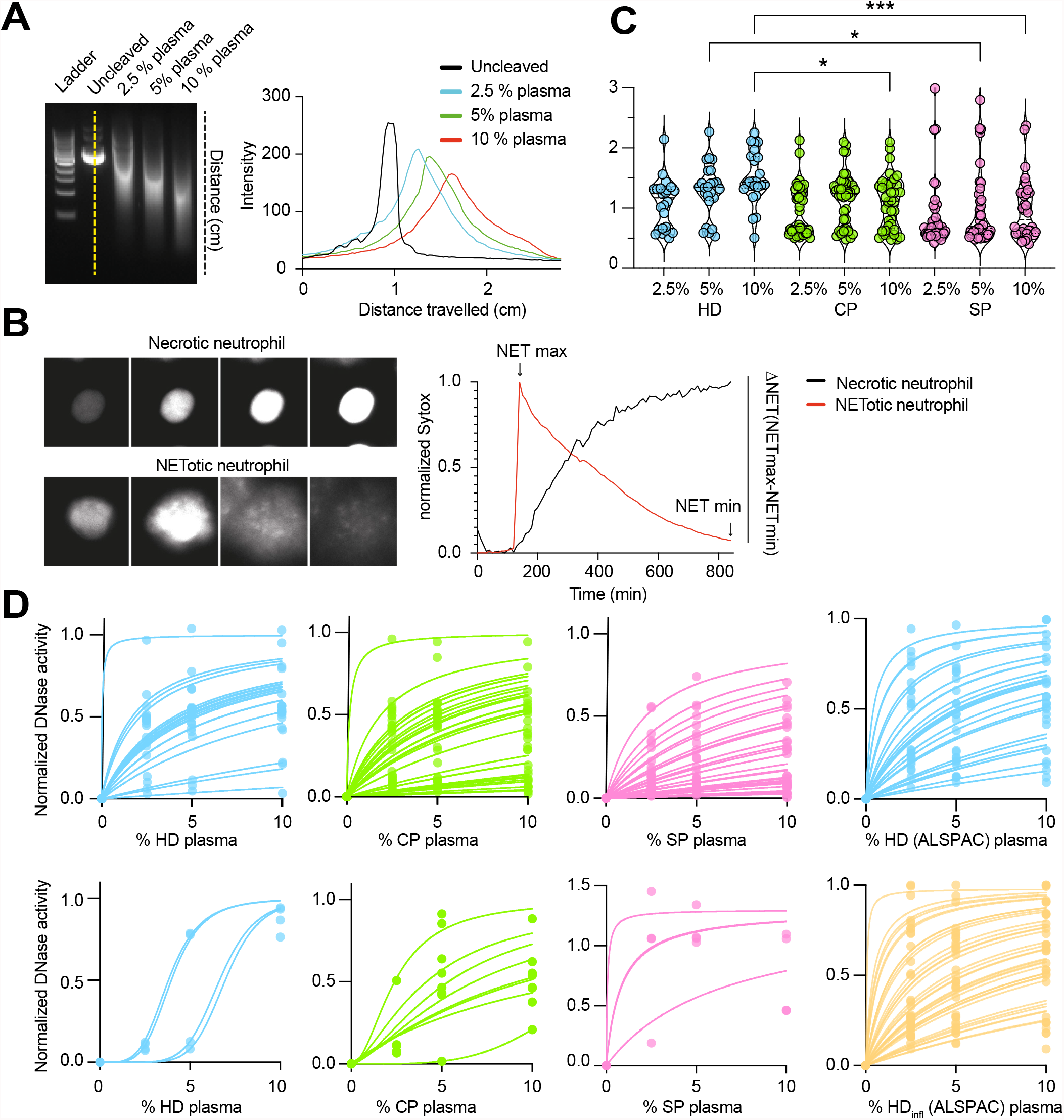
Quantification of DNase and NETase activities. **A**. Quantification of plasmid DNA degradation by different dilutions of human plasma. Each agarose gel electrophoresis lane was digitally quantified to locate the DNA signal peak and measure the distance travelled (cm). **B**. Human neutrophils activated by phorbol myristate acetate (PMA) in the presence of 2% plasma were monitored by time-lapse microscopy for NET degradation measurements and each neutrophil was identified and its mean fluorescence tracked over 8 hrs. Necrotic neutrophils were distinguished from NET-forming neutrophils by the pattern of change in fluorescence (right). In NET forming neutrophils, NETs dissolved over time leading to a gradual decrease in fluorescence. The fraction of fluorescence lost over time was used to calculate the mean NETase activity of the plasma against thousands of NETs. **C**. DNase activity measured as in (A) in healthy donor (HD), SARS-CoV-2 infected patients (CP) and microbial sepsis (SP) plasmas at 2.5%, 5% and 10% dilution. Statistical analysis by one-way Anova test. **D**. Best fitting curves for DNase activity values per donor in (C), as well as HD and HD_infl_ from the ALPAC study used to calculate the half-maximal activity dilution (D_50_). Curves fitted by non-linear regression to a three parameter association function.

**Supplemental Figure 3.**
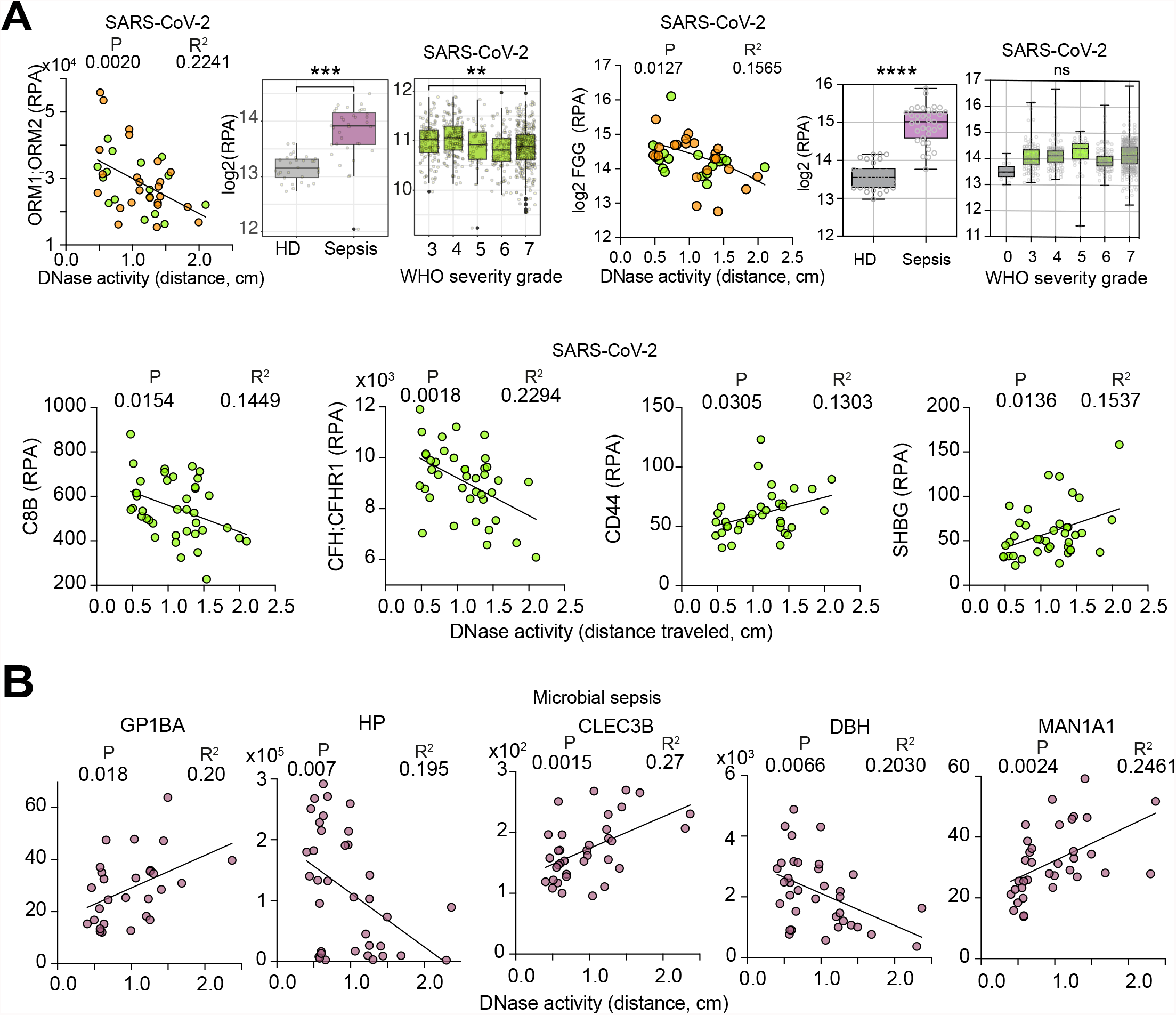
Correlations between DNA degradation activity and plasma proteomes. **A**. DNase activity measured as the distance travelled during gel electrophoresis by the substrate DNA after digestion with a 10% dilution of plasma from 40 WHO-7 SARS-CoV-2 infected patients (24 WHO-7, 3 WHO-3 and 13 WHO-4 severity grade samples). Significant correlations between DNase activity in 10% plasma and plasma proteins measured by mass spectrometry. (Middle panel) Differences between healthy donor (HD) and sepsis patient plasmas (SP), as well as protein levels in 687 CP plasma samples segregated by the associated WHO severity grade are shown for comparison (right panel). **B**. Significant correlations between DNase activity and plasma proteins measured by mass spectrometry in 36 microbial sepsis patient plasma samples. Statistical analysis by Mann-Whitney for single comparisons or one-way Anova test for multiple comparisons, and simple linear regression fitting where P values appear on the left and R^2^ values on the right. * 0.05-0.01, ** 0.01-0.001, *** 0.001-0.0001 and **** <0.0001.

**Supplemental Figure 4.**
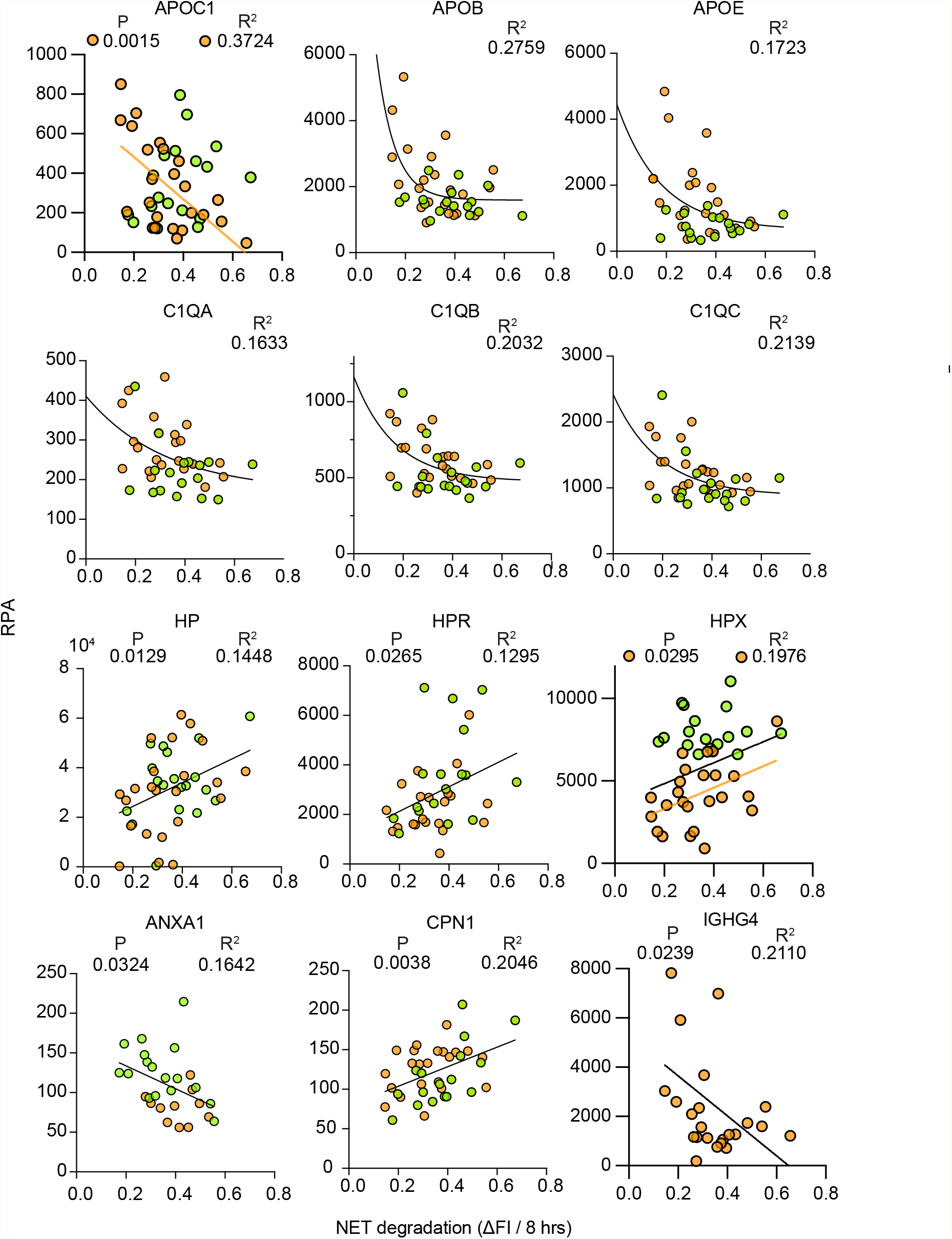
Correlations between NET degradation activity and plasma proteomes. Significant correlations between NETase activity and plasma proteins measured by mass spectrometry in 42 SARS-CoV-2 patient plasma samples. Proteins were identified by proteome-wide corelation analysis for significant correlation by simple linear regression, gaussian or variable decay fitting. P values appear on the left and R^2^ values on the right.

**Supplemental Figure 5.**
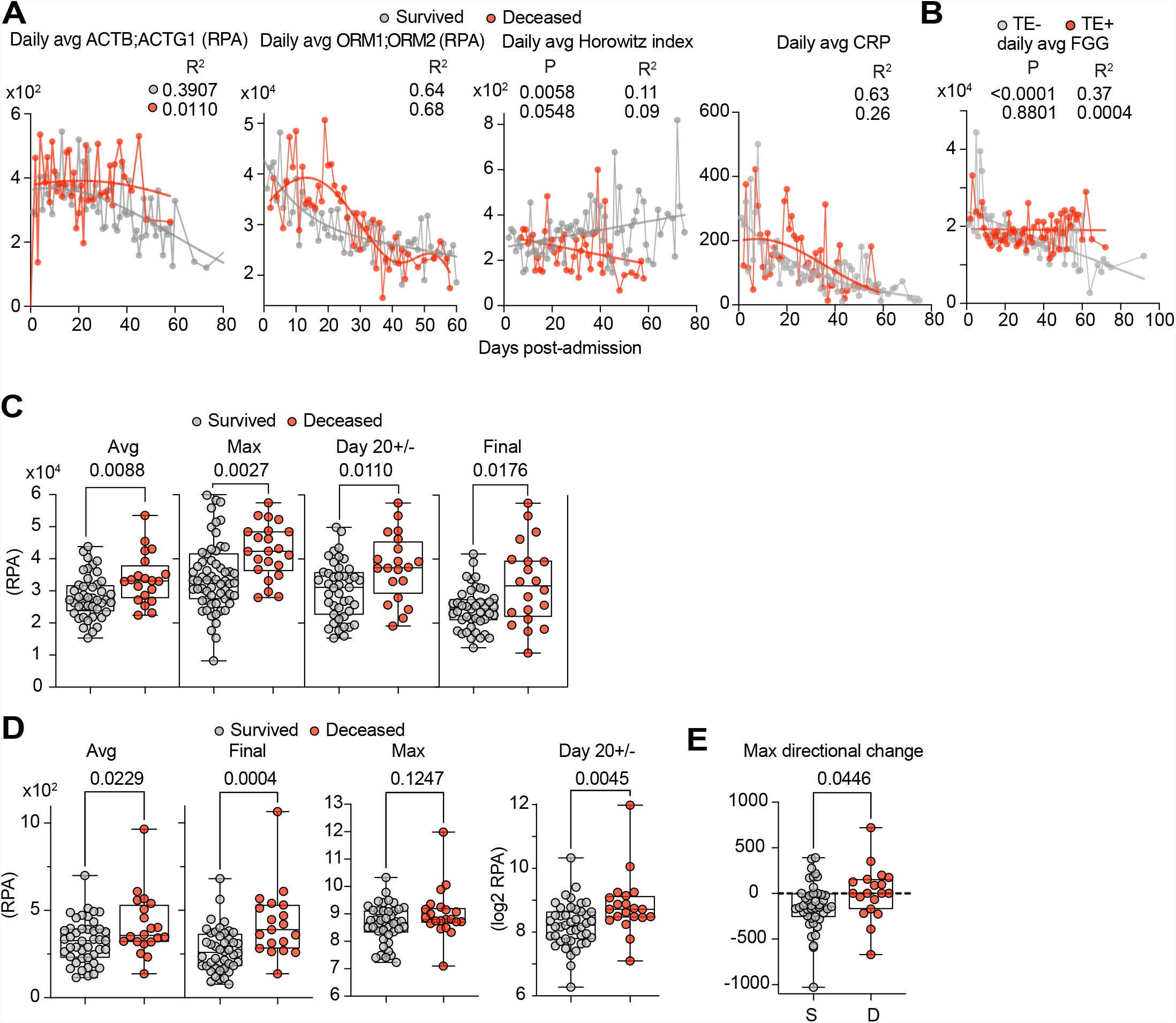
Dynamics of actin, ORM proteins, and clinical parameters during hospitalization. **A**. Daily cumulative average values relative to time post-admission for actin (ACTB;ACTG1), ORM1;ORM2, Horowitz index and CRP measured by mass spectrometry in the plasmas of 63 SARS-CoV-2 patients that reached a maximum WHO severity grade of 7, separated by survival outcome: patients that survived (grey) or deceased (red). **B**. Daily cumulative average FGG levels in patients seggregated by whether they presented with thromboembolism (TE). **C** and **D**. Average, maximum, D20+/- (reading from sample closest to day 20 post-admission) and the final plasma sample ORM1;OMR2 (c) and actin (ACTB;ACTG1) (d) readings per patient grouped by survival outcome. **E**. The directional difference between the maximum and minimum longitudinal ACTB;ACTG1 values per patient grouped by survival outcome. Positive values represent increases over time, whereas negative values indicate a reduction. Statistical analysis by Mann-Whitney test for single comparisons, Mantel-Cox test for survival, simple linear regression, or non-linear exponential or Gaussian fitting where P values appear on the left and R^2^ values on the right.

**Supplemental Figure 6.**
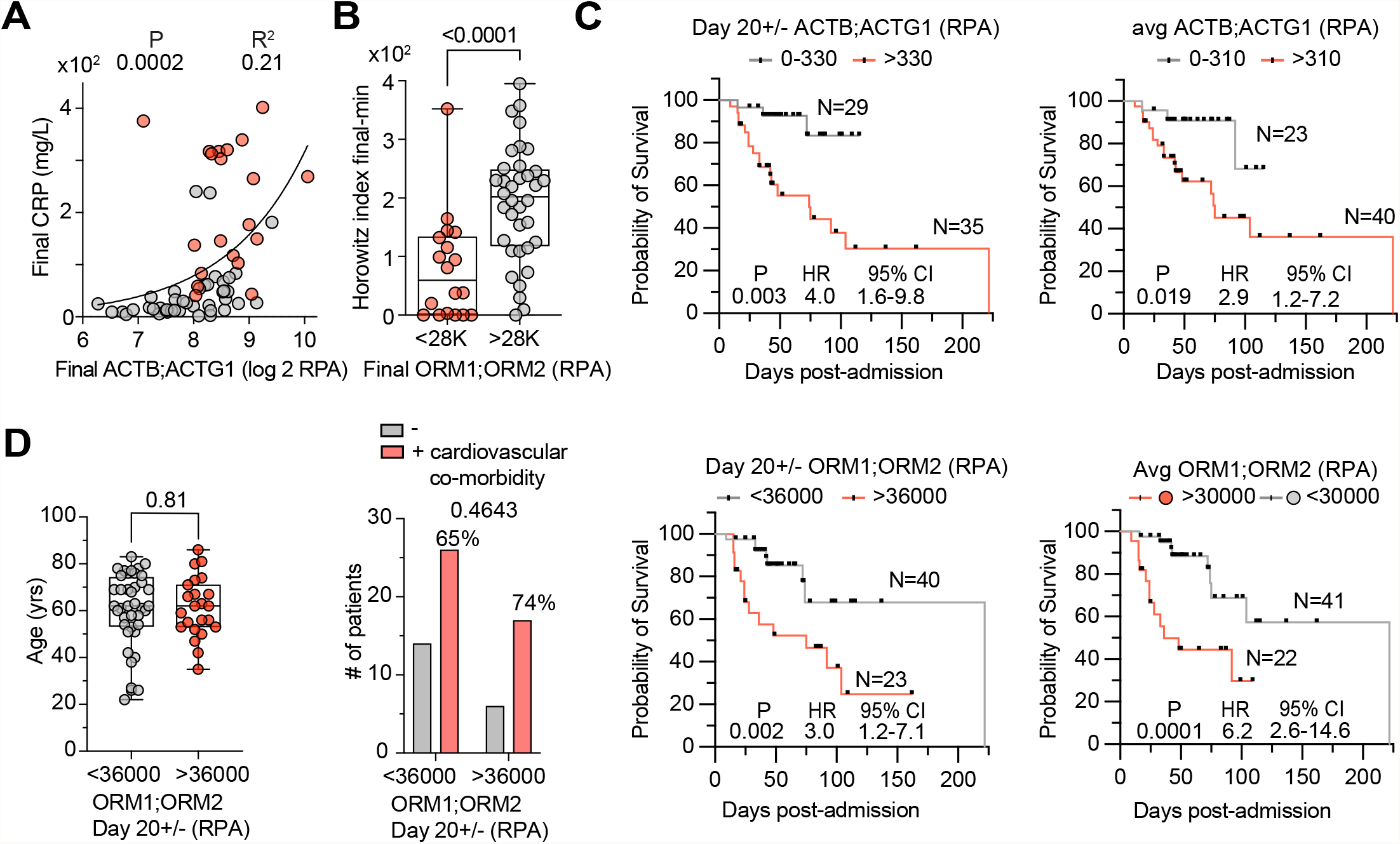
Relationship between actin, ORM proteins and FGG in the plasma of SARS-CoV-2 infected patients. Analysis of 63 SARS-CoV-2 patients that reached WHO severity grade 7: **A**. Correlation between the final actin and plasma levels of CRP. **B**. Difference between the final and minimum Horowitz index values per patient seggregated by the whether the final ORM protein values for each patient were above or below 28000 RPA. **C**. Probability of survival in patients clustered into groups according to either their proximal to day 20 (Day 20+/-) or longitudinal average ORM1:ORM2 or ACTB;ACTG1 values per patient. Black bars indicate censoring events. **D**. Age and incidence of cardiovascular co-morbidities in patienst grouped according to their D20+/- ORM protein levels. Statistical analysis by Mann-Whitney test for single comparisons, simple linear regression for fitting or Fisher’s exact test for contigency distribution analysis. P values appear on the left and R^2^ values on the right. Survival probabilities clalculated by Mantel-Cox survival analysis.

**Supplemental Figure 7.**
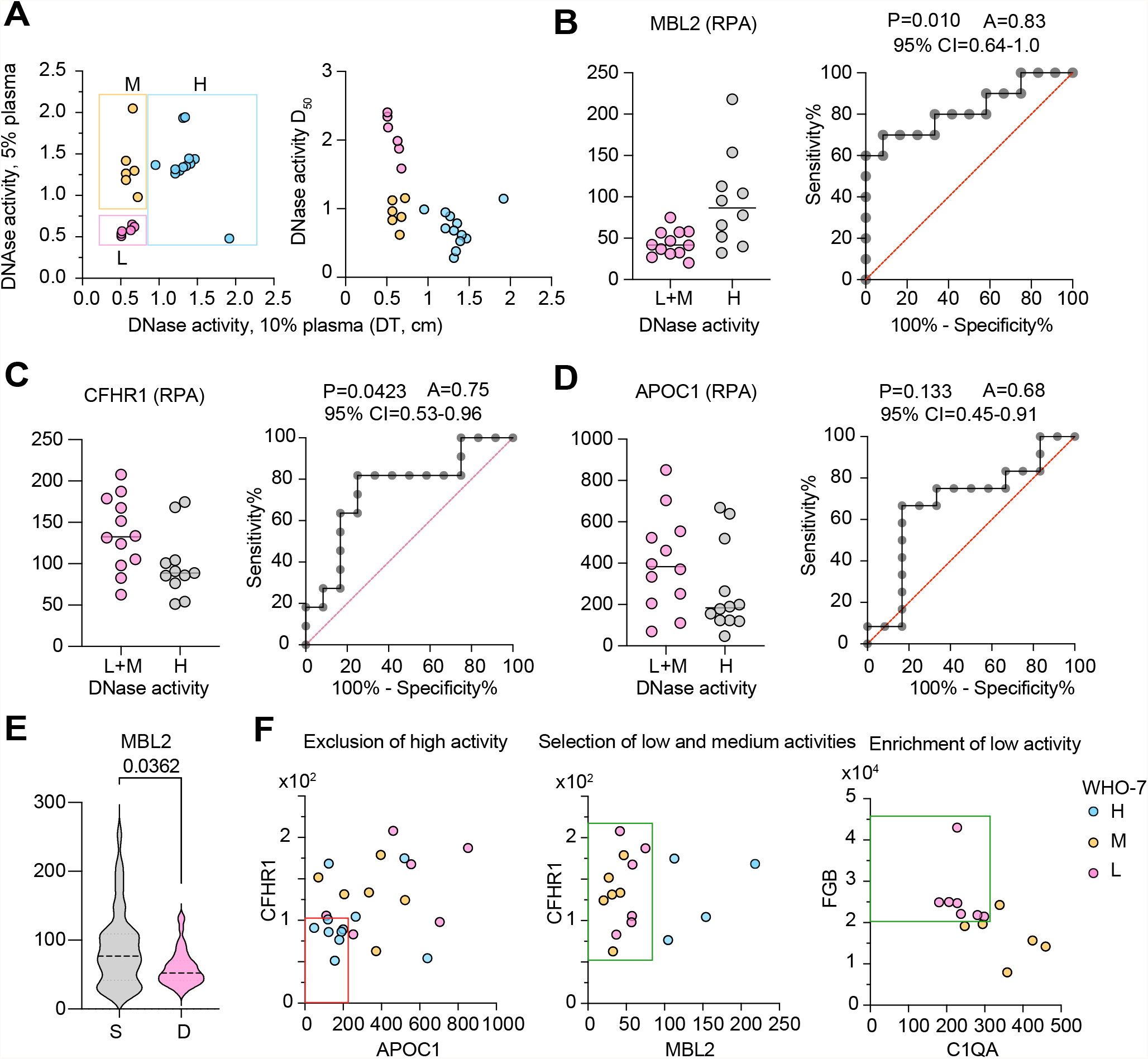
Gating strategies for DNase activity profiling in SARS-CoV-2 infected patient plasma samples. **A**. DNase activity measured as the distance travelled during gel electrophoresis by the substrate DNA after digestion with 2.5 or 5% dilutions of plasma from 24 WHO-7 SARS-CoV-2 infected patients. Relationship of 2.5% plasma degradation and D_50_ (right panel). Boxes show samples grouped by activity where L is low, M is medium and H is high activity. **B-D**. MBL2, CFHR1 and APOC1 levels in low+medium and high DNase activity samples and area under the curve (AUC) plot. **E**. MBL2 levels in 300 samples from 52 SARS-CoV-2 infected patients with maximum WHO severity grade 7 segregated into survivors (S) and deceased (D). **F**. Gating strategy for assignment of low, medium, high DNase activity profiles based on the 24 tested WHO-7 CP plasma samples. CFHR1^low^;APOC1^low^ samples were excluded as DNase high (red box). The remaining DNase high samples were exluded as MBL2^high^. Low and medium DNase activity samples were separated using FGB and C1QA protein levels.

**Supplemental Figure 8.**
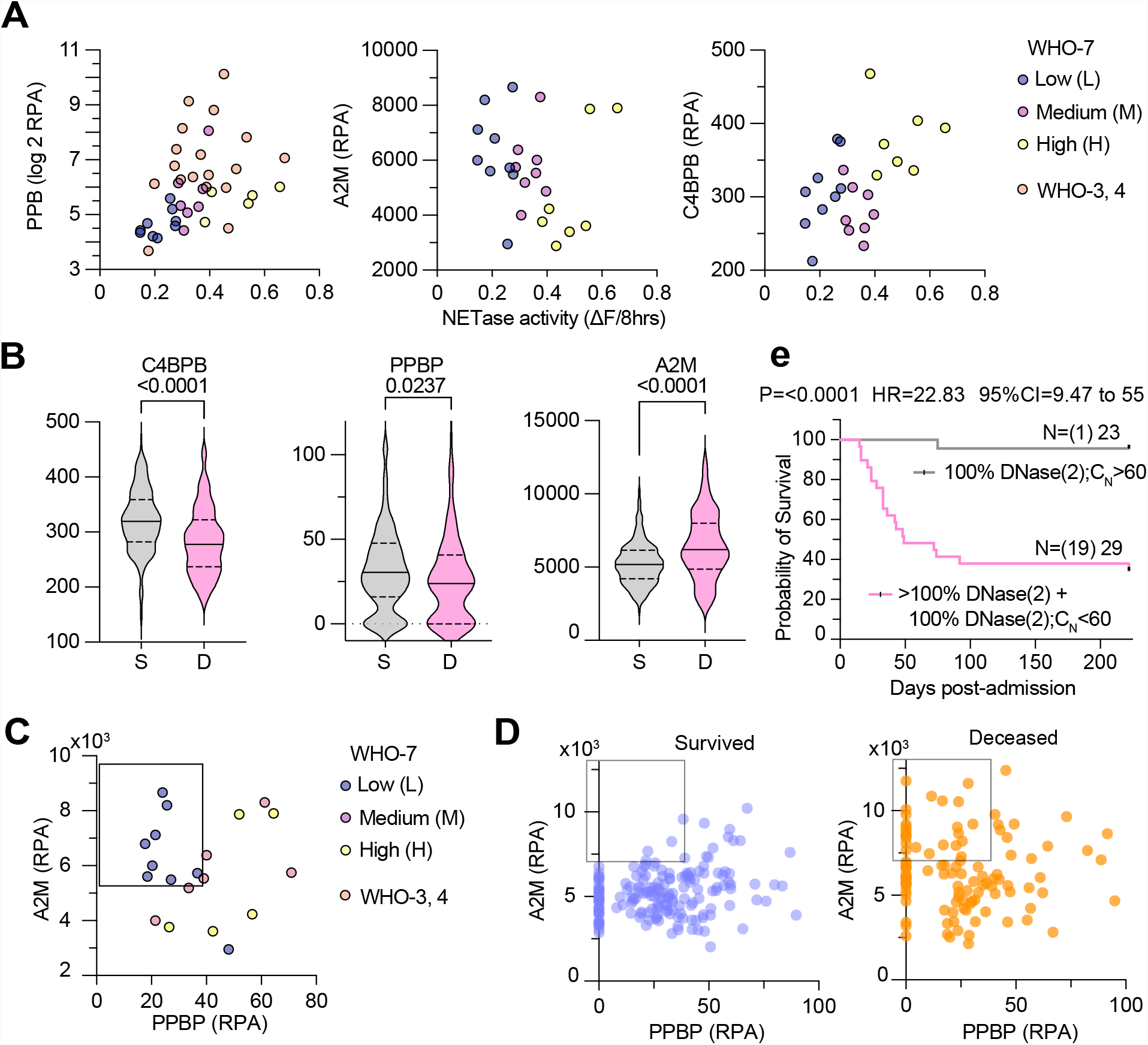
Gating strategies for NETase activity profiling in SARS-CoV-2 infected patient plasma samples. **A**. Correlation between NETase activity and PPBP, A2M or C4BPB in 24 tested WHO-7 SARS-CoV-2 infected patient plasma samples. Color scheme denotes samples with different activity ranges: L for low, M for medium and H for high NETase activity. **B**. Plots for C4BPB, PPBP, A2M values in 300 WHO-7 severity grade plasma samples from 52 surviving (S) and deceased (D) SARS-CoV-2 infected patients. **C**. A2M and PPBP plots depicting the criteria used to define low NETase activity (0) samples as PBPB<40;A2M>5500 RPA. DNase high samples were identified as samples that exceed C4BPB>330 RPA. The remaining samples were assigned to NETase intermediate activity. **D**. 300 WHO-7 severity grade plasma samples from 52 surviving (S) and deceased (D) SARS-CoV-2 infected patients plotted for A2M and PPBP. The box indicates the difference in the number of low NETase profiled samples associated with the two groups. **E**. Survival of patients with combined C_N_ above 60 and at least 80% high DNase activity sample content or a C_N_ below 60 and lower than 80% high DNase activity sample content, assuming that all censored patients survived after being discharged. Numbers in parentheses denote the number of deaths per group. Statistical analysis by Man-Whitney test for single comparisons and Mantel-Cox survival analysis.

**Supplemental Figure 9.**
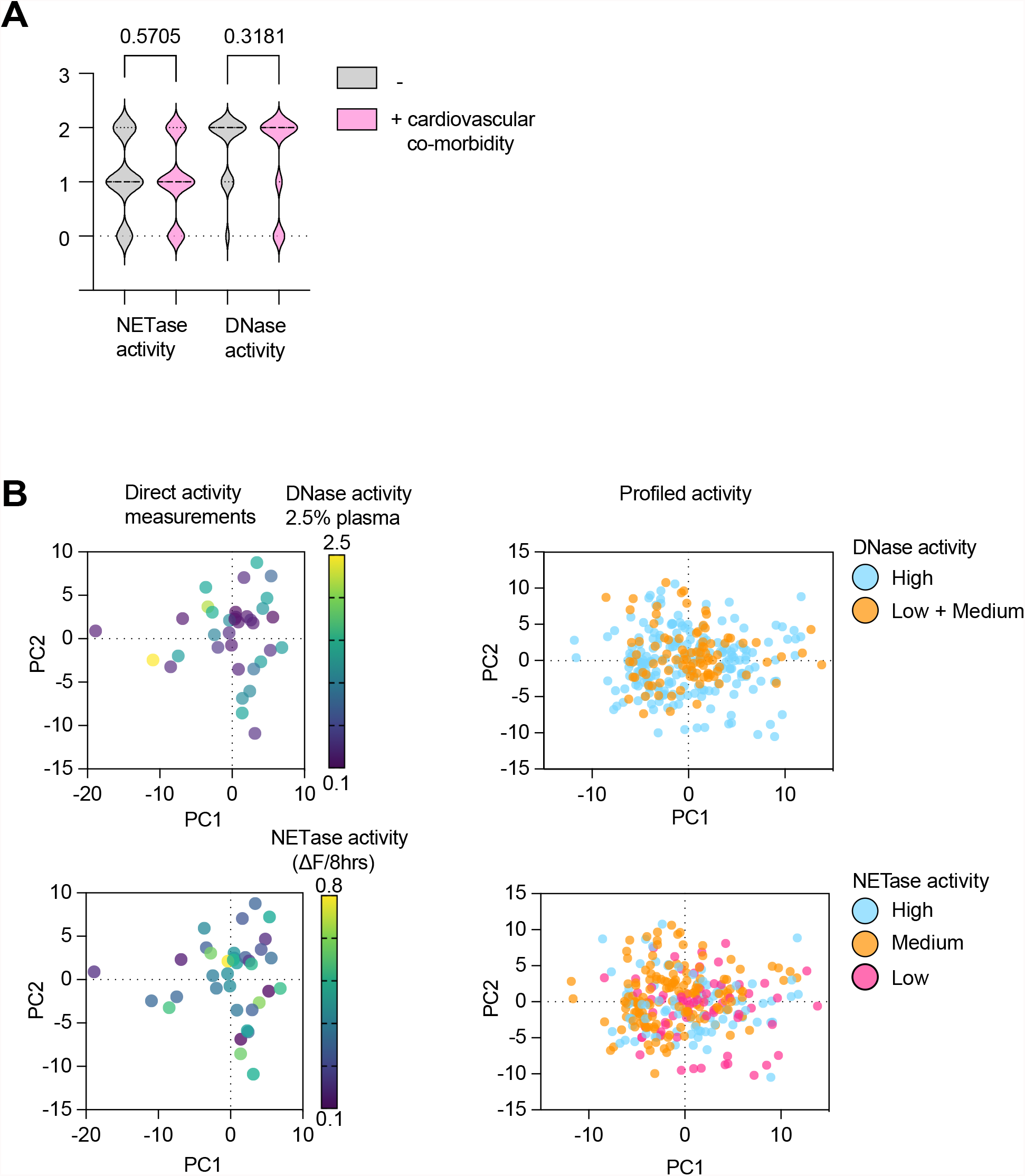
Distribution of activities relative to cardiovascular co-morbidities and PCA analysis of SARS-CoV-2 infected patient plasma proteomes. **A**. Incidence of profiled DNase and NETase activity samples from patients with and without cardiovascular co-morbidities. **B**. PCA analysis of the proteomes of the samples measured directly (left panels) and the total 300 WHO-7 samples from 52 SARS-CoV-2 infected patients (right panels) labeleld by their measured or profiled DNase (upper panels) and NETase (lower panels) activities. Different colors denote different ranges of activity. Statistical analysis by Man-Whitney test for single comparisons.

